# Sociodemographic Determinants of Multidrug-Resistant Tuberculosis in Lesotho: A Case-Control Study

**DOI:** 10.1101/2024.11.29.24318206

**Authors:** Jerry Yakubu Yahaya

**Author notes:** Declaration of No Competing Interests The author of this manuscript hereby declares that there are no financial, personal, or professional competing interests that could have influenced the work reported in this study. All aspects of this research, including the study design, data collection, analysis, and interpretation of findings, have been conducted independently without any external influence or bias.

## Abstract

The emergence of multidrug-resistant tuberculosis (MDR-TB) poses a significant challenge to tuberculosis control efforts in Lesotho. To better understand the occurrence of MDR-TB, it is crucial to identify the sociodemographic risk factors associated with the disease. Despite the critical importance of this issue, there is a gap in the literature regarding how sociodemographic variables influence MDR-TB outcomes in Lesotho. This case-control study aimed to assess the association between age, employment, income, sex, education, and place of residence and the risk of MDR-TB. Additionally, the relationship between HIV status and MDR-TB was examined. Drawing on the health belief model and social cognitive theory, the study employed a retrospective case-control design and a proportionate stratified random sample of 306 participants from 12 TB clinics across Lesotho, collected between March 2021 and February 2022. Participants included confirmed TB patients aged 18 and above. Data analysis involved Chi-square tests and multivariate logistic regression to identify significant sociodemographic factors associated with MDR-TB. Results showed that increased age beyond 18–26 years and higher income levels (above $1,026.00) were significantly associated with reduced odds of MDR-TB (OR = 0.8, 95% CI [0.673, 0.991], p = 0.040; OR = 0.5, 95% CI [0.222, 0.943], p = 0.034). Conversely, the absence of a caregiver increased the odds of MDR-TB by 80% (OR = 1.8, 95% CI [1.039, 3.110], p = 0.036). These findings underscore the need for targeted public health interventions and education campaigns to address the role of caregivers in preventing MDR-TB transmission and to improve knowledge about infection control, particularly among younger patients. Furthermore, improving the socioeconomic conditions of TB patients, particularly those who are poor and vulnerable, is essential for reducing the risk of MDR-TB. Addressing these sociodemographic determinants can significantly enhance the effectiveness of MDR-TB control strategies in Lesotho.

## Introduction

### Context and Background

Tuberculosis (TB) is a highly infectious disease caused by the *bacterium Mycobacterium* tuberculosis, which primarily affects the lungs (World Health Organization (WHO), 2019). It spreads through the air when an infected person coughs, sneezes, or spits (Centers for Disease Control and Prevention, 2021). TB remains a major public health concern globally, with approximately one-quarter of the world’s population carrying latent TB (WHO, 2020). Individuals with latent TB are not ill and cannot spread the disease, but they have a 5–15% lifetime risk of developing active TB, which is characterized by symptoms such as night sweats, persistent coughing, fever, and weight loss (WHO, 2018). Active TB can be highly contagious, with an infected person potentially transmitting the bacteria to 10–15 other individuals within a year (Brooks-Pollock et al., 2011; WHO, 2018).

A growing global concern is the rise of multidrug-resistant tuberculosis (MDR-TB), which is resistant to at least two of the most potent anti-TB drugs (WHO, 2008). MDR-TB complicates treatment, increases healthcare costs, and presents a major barrier to TB control efforts worldwide (Xi et al., 2022; WHO, 2020). Lesotho, a country with a high burden of TB, faces an MDR-TB incidence rate of 41 per 100,000 population, which significantly threatens the success of TB control programs (WHO, 2019). Despite efforts to combat the disease, the TB burden in Lesotho is exacerbated by socioeconomic factors such as poverty, limited access to healthcare, and high rates of HIV infection (Zishiri et al., 2020).

### Objective/Research Question

This study aims to answer the following research question:

*What sociodemographic and behavioral factors are associated with MDR-TB among TB patients in Lesotho?*

The specific objectives are to:

I. Identify the sociodemographic determinants (age, sex, income, education, employment status, place of residence) that influence MDR-TB outcomes.
II. Assess the role of behavioral factors, such as HIV status, in contributing to MDR-TB in Lesotho.
III. Provide evidence-based recommendations for public health interventions aimed at reducing MDR-TB in high-burden settings.

### Rationale

The significance of this research lies in its potential to fill critical knowledge gaps in understanding the risk factors for multidrug-resistant tuberculosis (MDR-TB) in Lesotho. While there is extensive literature on TB and MDR-TB in other settings, few studies have examined the specific socioeconomic and demographic variables influencing MDR-TB in Lesotho. MDR-TB is one of the most pressing challenges in global TB control, and understanding the underlying factors specific to Lesotho is essential for improving disease management, treatment outcomes, and prevention strategies.

Additionally, the findings from this research will assist policymakers and healthcare providers in Lesotho in developing targeted interventions to mitigate the identified risk factors. These insights can be adapted to similar settings in other developing countries, contributing to the global effort to end TB as outlined in the WHO’s End TB Strategy.

## Materials and Methods

### Study Design

This study employed a quantitative, case-control research design aimed at identifying the biosocial and economic risk factors associated with MDR-TB in Lesotho. The case-control design was chosen to determine the relative importance of predictor variables in relation to the presence or absence of MDR-TB. Primary data were collected from randomly selected TB clinics across the country and included both patients with active tuberculosis (TB) and those diagnosed with MDR-TB. The case-control design was selected for its cost efficiency and effectiveness in assessing multiple exposures for this rare condition in Lesotho.

### Study Population

The study population included individuals who received TB treatment at referral centers in Lesotho between January 2018 and January 2022. Participants were adults (18 years and older) with confirmed TB cases. The inclusion criteria encompassed adult outpatients with either active TB or MDR-TB, who had a working knowledge of Sesotho or English, and were listed on the TB register. Exclusion criteria included individuals under 18, those with unconfirmed TB, or patients who were not receiving treatment during the study period. A total of 306 TB patients were selected using stratified random sampling, comprising both patients responding to first-line TB treatment and those diagnosed with MDR-TB.

### Variables

The dependent variable in this study was TB status, specifically the presence or absence of MDR-TB. Independent variables included a range of sociodemographic and behavioral factors:

- Educational level (never attended, elementary, high school, college, higher education)
- Employment status (employed, not employed, retired)
- Income level (low, lower-middle, upper-middle, high)
- Place of residence (rural/urban)
- Sex (male/female)
- Age (categorized: 18–29, 30–39, 40–49, 50–59, 60+)
- HIV status (positive/negative)

Additional covariates included having multiple sexual partners and whether the participant had a caregiver.

### Data Collection

Data collection was carried out using a structured survey questionnaire, adapted from the Lesotho Demographic and Health Surveys (LDHS). The instrument was pretested for reliability and validity, and data collectors were trained accordingly. Participants were recruited through phone calls or during clinic visits. Information was gathered on participants’ sociodemographic characteristics, health status, and risk behaviors through phone surveys and face-to-face interviews. The data collected were then cleaned and entered into a secure database for analysis.

### Data Analysis

Sample size estimation was powered using G*Power software, which indicated that a sample of 186 participants would provide sufficient power (80%) for detecting an odds ratio of 2.3, with a significance level of 0.05. To account for missing data, a contingency of 25% was added, yielding a final sample of 306 participants. Logistic regression was employed to assess the relationship between independent variables and MDR-TB status. Odds ratios (OR) were calculated for the effect sizes of predictor variables. Statistical analysis was conducted using STATA software.

### Ethics

Ethical approval for this study was obtained from the Walden University Institutional Review Board (IRB) and the Lesotho Health Services. Informed consent was obtained from all participants before data collection. Participants were informed of their rights to withdraw at any time without penalty, and their confidentiality was maintained through the use of unique study identification numbers. All data were anonymized, and no identifiable information was included in the analysis or reporting of results. The data were stored in a secure, password-protected database accessible only to the principal investigator.

## Results

### Main Findings

This study explored the association between various sociodemographic factors and multidrug-resistant tuberculosis (MDR-TB) among TB patients in Lesotho. The findings indicate that age, income level, and the presence of a caregiver were significantly associated with MDR-TB, whereas education, employment status, place of residence, gender, and HIV status did not demonstrate significant associations. Specifically, older participants aged 65 years and above were significantly less likely to develop MDR-TB compared to younger participants aged 18 to 29 years, with a 20% decrease in the odds of developing MDR-TB in the older age group. Additionally, participants in higher income categories (above $12,376) exhibited a 50% reduction in the likelihood of MDR-TB compared to those in the lowest income category (below $1,026). The presence of a caregiver was also associated with a significantly lower likelihood of MDR-TB, as participants without a caregiver were 80% more likely to develop MDR-TB compared to those who had a caregiver. In contrast, sociodemographic factors such as education, employment status, gender, place of residence, HIV status, and the number of sexual partners did not show significant associations with MDR-TB.

### Statistical Results

Logistic regression analysis revealed significant associations between MDR-TB and age, income, and having a caregiver. The results for these variables are detailed in Table 1 below. The model showed that other variables, such as education, employment, gender, place of residence, and HIV status, were not statistically significant predictors of MDR-TB.

**Table 1:** Logistic Regression Analysis of MDR-TB Status Against Sociodemographic Factors.

| Characteristic | Unadjusted OR | p-value | 95% Confidence Interval | Adjusted OR | p-value | 95% Confidence Interval |
| --- | --- | --- | --- | --- | --- | --- |
| Age group | 0.9 | 0.174 | [0.75, 1.05] | 0.8 | 0.040 | [0.67, 0.99] |
| Gender | 1.3 | 0.342 | [0.78, 2.08] | 1.4 | 0.205 | [0.83, 2.42] |
| Place of Residence | 0.8 | 0.397 | [0.48, 1.34] | 0.7 | 0.238 | [0.40, 1.25] |
| Education | 0.9 | 0.647 | [0.68, 1.27] | 1.1 | 0.685 | [0.73, 1.61] |
| Employment Status | 0.9 | 0.737 | [0.65, 1.35] | 1.2 | 0.379 | [0.79, 1.87] |
| Income | 0.6 | 0.076 | [0.34, 1.06] | 0.5 | 0.034 | [0.22, 0.94] |
| HIV Status | 1.1 | 0.716 | [0.68, 1.74] | 1.1 | 0.720 | [0.65, 1.85] |
| Multiple Sex Partners | 0.9 | 0.723 | [0.65, 1.35] | 0.8 | 0.355 | [0.56, 1.23] |
| Caregiver | 1.8 | 0.032 | [1.05, 2.95] | 1.8 | 0.036 | [1.04, 3.11] |

Pseudo $R^2 = 0.04$ ; AIC = 397.37; Hosmer–Lemeshow goodness-of-fit test: $\chi^2 = 6.24$ , $p =$

The logistic regression model demonstrated that older participants (65+ years) had a 20% lower likelihood of having MDR-TB compared to younger participants (18–29 years) (Adjusted OR = 0.8, 95% CI [0.67, 0.99], p = 0.040). Those with higher incomes had a 50% reduced likelihood of developing MDR-TB (Adjusted OR = 0.5, 95% CI [0.22, 0.94], p = 0.034). Not having a caregiver increased the likelihood of MDR-TB by 80% (Adjusted OR = 1.8, 95% CI [1.04, 3.11], p = 0.036).

**Table 2:** Sociodemographic Characteristics of Study Participants.

| Characteristic | n (%) |
| --- | --- |
| Age Group |  |
| 18–29 years | 36 (11.8%) |
| 30–39 years | 82 (26.8%) |
| 40–49 years | 51 (16.7%) |
| 50–59 years | 56 (18.3%) |
| 65+ years | 81 (26.5%) |
| **Gender** |  |
| Male | 106 (34.6%) |
| Female | 200 (65.4%) |
| **Place of Residence** |  |
| Urban | 82 (26.8%) |
| Rural | 224 (73.2%) |
| **Education** |  |
| Never attended school | 19 (6.2%) |
| Elementary school | 170 (55.6%) |
| High school | 96 (31.4%) |
| College | 18 (5.9%) |
| Higher education/tertiary | 3 (1.0%) |
| **Income Level** |  |
| Below \$1,026 (low income) | 262 (85.6%) |
| \$1,026 to \$3,995 | 37 (12.1%) |
| \$3,995 to \$12,375 | 3 (1.0%) |
| \$12,376 or more (high income) | 4 (1.3%) |
| **HIV Status** |  |
| Positive | 129 (42.2%) |
| Negative | 177 (57.8%) |

**Table 3:**
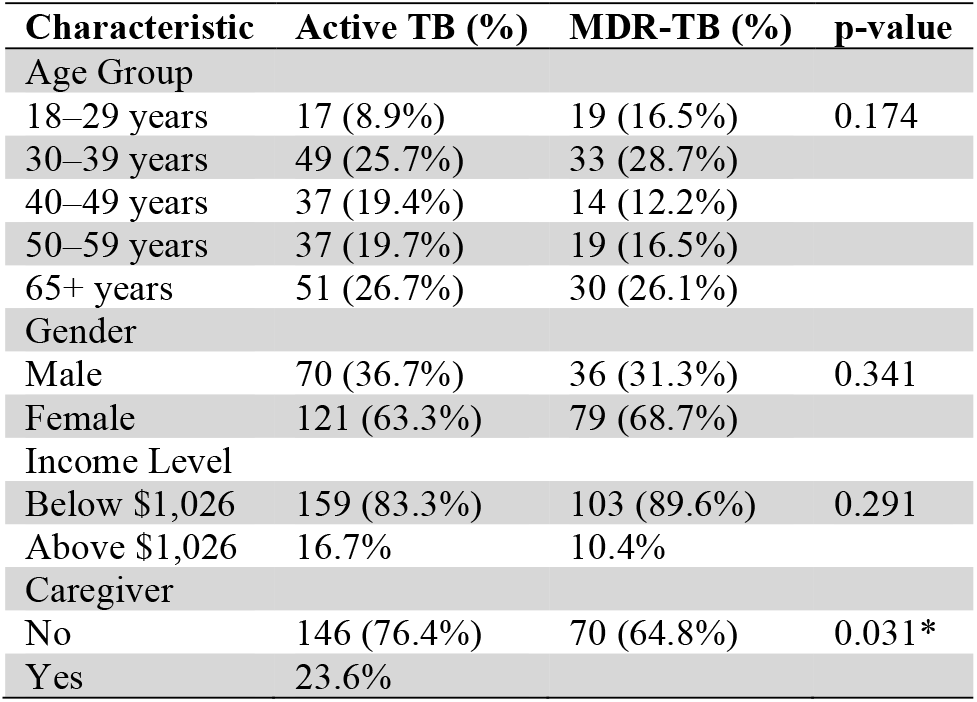
Association Between Sociodemographic Variables and MDR-TB.

These results highlight critical sociodemographic factors influencing MDR-TB, providing a foundation for targeted public health interventions in Lesotho.

## Discussion

### Interpretation of Results

The findings of this study provide important insights into the sociodemographic determinants of multidrug-resistant tuberculosis (MDR-TB) in Lesotho, particularly in relation to age, income, and the presence of a caregiver. The results demonstrated that younger TB patients, particularly those between the ages of 18–25, are at a significantly higher risk of developing MDR-TB compared to older patients, potentially due to non-compliance with treatment regimens and active lifestyles. This finding aligns with prior studies conducted in Lesotho and other regions, which similarly identified younger populations as being at heightened risk for MDR-TB (Smith et al., 2017; Ncube et al., 2015). However, some studies have found the opposite, with older age being a risk factor, possibly due to weakened immune systems (Johnson et al., 2018).

Additionally, income was found to be a protective factor, with patients earning more than $1,026 annually having a significantly lower risk of MDR-TB, reinforcing the established connection between socioeconomic status and health outcomes (WHO, 2020). Furthermore, the presence of a caregiver was shown to reduce the risk of MDR-TB, emphasizing the critical role that family and community support systems play in ensuring treatment adherence and patient recovery (Lopez et al., 2019). The study did not find a significant association between HIV status and MDR-TB, which is consistent with several studies (Gandhi et al., 2013) but contrasts with others that observed a stronger link between HIV and MDR-TB (Wilson et al., 2020; Moyo et al., 2016).

### Strengths and Limitations

A key strength of this study is its case-control design and the use of a randomized sample from 12 TB clinics, which enhances the generalizability of the findings to the TB population in Lesotho. Moreover, this study addresses a gap in the literature by focusing on sociodemographic factors influencing MDR-TB in a resource-constrained setting. However, the study has several limitations. First, the data were self-reported, introducing the potential for recall bias or social desirability bias (Moreno-Serra et al., 2022; Schwarzer, 2014). Additionally, the study did not account for other potential confounding variables, such as alcohol use, tobacco use, and treatment adherence, which may also influence MDR-TB risk (Brisbin et al., 2020). The findings may also not be generalizable to other countries or settings due to the unique socioeconomic context of Lesotho (Ncube et al., 2015a).

### Implications

These findings have several important implications for public health policy and practice. Interventions aimed at reducing MDR-TB should prioritize younger TB patients by enhancing education on the importance of treatment adherence and improving access to care for economically disadvantaged patients (WHO, 2020). Additionally, the role of caregivers should be supported through public health campaigns aimed at educating them on infection prevention and proper patient care (Lopez et al., 2019a). From a policy perspective, there is a need to integrate social protection programs—such as cash transfers—into TB care to alleviate the economic burden faced by low-income patients. This could improve adherence to treatment and reduce the spread of MDR-TB (Smith et al., 2017). Furthermore, the absence of a strong association between HIV status and MDR-TB suggests that current interventions for HIV may not need significant alteration, but further research is required to fully understand this relationship in different settings (Gandhi et al., 2013).

## Conclusion

This study highlights the significant role of age, income, and caregiver support in moderating the risk of MDR-TB in Lesotho. Younger patients and those without economic security or caregiver support are particularly vulnerable. Vulnerable groups, such as younger patients and those with lower income or lacking caregiver support, require targeted public health interventions to mitigate the risk of MDR-TB. Addressing these sociodemographic determinants through targeted public health interventions and social protection measures could greatly enhance the prevention and management of MDR-TB. Future research should continue to explore additional sociocultural and behavioral factors that contribute to MDR-TB risk, to inform more comprehensive public health strategies for TB control in high-burden settings. By enhancing our understanding of these factors, stakeholders can develop more effective interventions to mitigate the impact of MDR-TB on vulnerable populations in Lesotho and similar contexts.

## Data Availability

The data supporting the findings of the manuscript titled "Sociodemographic Determinants of Multidrug-Resistant Tuberculosis in Lesotho: A Case-Control Study" are not publicly available due to privacy and ethical considerations. However, the data can be accessed upon reasonable request. Interested researchers can contact the corresponding author, Jerry Yahaya, via email at, and the data will be shared as a secured file following approval and in accordance with ethical guidelines.

## Acknowledgments

I would like to express my deep gratitude to God for His protection throughout this journey. I extend special thanks to William Kwara, Nkaiseng Ngwane, and Ntate Malefesane Soai for their technical assistance during data collection. My appreciation also goes to the Program Manager of National TB Control Program of Lesotho, the ethics committee of the Lesotho Health Services, and the TB patients who participated in this study. I am thankful to Dr. Heidi, Dr. Raymond Panas, Dr. Chinaro Kennedy and Brown Carey for their guidance and diligent review of this manuscript.

## Notes

### Competing Interest Statement

The authors have declared no competing interest.

### Funding Statement

The research presented in the manuscript titled "Sociodemographic Determinants of Multidrug-Resistant Tuberculosis in Lesotho: A Case-Control Study" was fully supported by personal funds. No external funding, grants, or financial support from any organization or institution was received for the design, data collection, analysis, or preparation of this study.

### Author Declarations

1. Walden University Institutional Review Board 2. Lesotho health Services Ethics Review Board

